# Heterogeneity in testing for infectious diseases

**DOI:** 10.1101/2022.01.11.22269086

**Authors:** Christian Berrig, Viggo Andreasen, Bjarke Frost Nielsen

## Abstract

Testing strategies have varied widely between nation states during the COVID-19 pandemic, in intensity as well as methodology. Some countries have mainly performed diagnostic testing while others have opted for mass-screening for the presence of SARS-CoV-2 as well. COVID passport solutions have been introduced, in which access to several aspects of public life requires either testing, proof of vaccination or a combination thereof. This creates a coupling between personal activity levels and testing behaviour which, as we show, leverages the heterogeneous behaviours in the population and turns this heterogeneity from a disadvantage to an advantage for epidemic control.

## 1. Introduction

During the coronavirus disease 2019 (COVID-19) pandemic, population-wide as well as targeted mitigation strategies have played crucial roles in terms of keeping societies functional in the face of a highly transmissible novel pathogen. Non-pharmaceutical interventions (NPIs) such as lockdowns, mass testing and contact tracing have played a prominent role, due to the scale at which they have been deployed. This study focuses on regular screening programmes, a class of NPIs which has been employed by several nations as well as institutions and employers around the world. The strategy has been particularly predominant among European countries as well as in the United Arab Emirates [1–12], and picked up speed aided by the availability of relatively inexpensive rapid antigen tests [13, 14].

There have been enormous national and regional differences in the level of testing for SARS-CoV-2 and in the nature of testing programs themselves. This is partly to do with the different purposes of testing strategies. The primary overall purpose of testing is of course to identify cases of disease, but for transmissible diseases such as COVID-19, contact tracing and genetic surveillance of the pathogen are also common objectives.

*Diagnostic testing* seeks to confirm (or rule out) the presence of the pathogen in a person suspected of being ill, most often on the basis of having shown symptoms. *Screening* is less targeted and looks for infections across entire groups, where no symptoms are reported [15]. Mass-testing represents one extreme of this spectrum, wherein large swathes of the population are invited for screening. However, in the case of a large outbreak of a fast-moving pathogen such as SARS-CoV-2, mass-testing at a single point in time may not be sufficient and schemes of repeated or regular testing may be prudent. This is especially true when tests have less-than-ideal sensitivities, such as is the case for rapid lateral-flow antigen (AG) tests [16], which may be made up for by increasing the frequency of testing [17]. In this paper, we exclusively consider regular screening for an infectious disease such as COVID-19.

Theoretical analyses of testing schemes have generally assumed that participating populations were homogeneous when it comes to testing behaviour [18]. However, empirical evidence shows that testing frequencies often vary widely, even on an aggregate level [19–22]. The influence of within-population heterogeneity in testing frequency has not been thoroughly understood, and this is the main problem we tackle from a theoretical point of view with this article.

Adding further complexity, several nations have introduced COVID ‘passport’ systems in which testing (and/or immunity through vaccination or previous infection) is a requirement in order to participate in many parts of public life, such as dining out, visiting bars and nightclubs as well as going to concerts and other large events[23, 24] – in some cases even to go into work[25]. Our analysis focuses on the testing aspect in isolation, and so does not assume any particular immunity structure in the population. The testing requirement implemented through COVID passports introduces a coupling between activity levels (understood as epidemiologically relevant contact rates) and testing behaviour. Those who are highly active will generally need a valid COVID passport at any given time, meaning that they are likely to undergo regular testing 2-3 times per week.

We are thus presented with two main questions: How do heterogeneity in testing rates impact the efficiency of a regular testing scheme? And how do correlations between activity levels and test frequency affect this?

## 2. Methods

### Homogeneous testing

The mitigative effect of regular testing relies on decreasing the amount of contact time that an infected individual is likely to have during the infectious period. Just like in classic compartmental transmission models of the SIR-type [26, 27], we assume that each individual has a certain rate of transmission – a probability per unit time – while infectious.

We first develop the mathematical framework for regular testing in the homogeneous case in which each individual undergoes regular testing with the same frequency *f* and thus with a testing interval of 1*/f*. We let the length of the infectious period be unity, *T*_*I*_= 1, so that all times are measured in units of the infectious period and frequencies are measured in units of 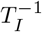. In the homogeneous case, the transmission rate is assumed to be identical for all infected individuals as well, such that the mitigation effect obtained by testing and subsequent isolation depends only on the time of the first positive test result for each individual.

In order to ascertain the reduction in infections due to regular testing, we must take into account the susceptibility *r*_*S*_ of the individual (referring here to the risk of becoming infected), as well as the infectiousness *r*_*I*_once infected (referring to the rate at which the disease is passed on). The total infectious burden that each individual contributes is proportional to the product *r*_*S*_ × *r*_*I*_ × *t*_*I*_ with *t*_*I*_ the time spent being infectious and non-isolated. The effect of regular testing is then to shorten the time that an infected individual is likely to spend outside of isolation.

The timing of the first test after the beginning of the infectious period is of course stochastic in such a regular testing scheme. We denote the probability density distribution of this time *P*_1_(*t*; *f*) where *t* ∈ ℝ_+_. Note that we do not restrict *t* to lie within the infectious period (*t* ≤ 1), since the first test may occur *after* the infectious period has elapsed if the frequency of testing is sufficiently low (*f* < 1). *P*_1_(*t*; *f*) is thus a ‘box-distribution’ given by:

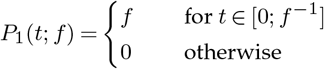

Once the time *t*_1_ of the first test has been determined, the following tests occur deterministically at times *t*_*n*_ = *t*_1_ + (*n* − 1)/*f*. The distribution of the time of the *n*’th test is thus found by summing over all the possible times of the first test:

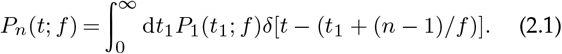

where we have made use of Dirac’s delta function which has the properties that

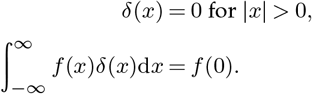

Suppose first that just one test can be performed during the infectious period. Once a test occurs, the probability of an infected individual being detected is then given by the test sensitivity *s* ≤ 1. If a detection takes place, the fractional reduction in reproductive number is of course 1 − *t*. The expected reduction is thus:

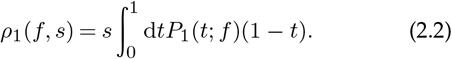

Note that the integration limits ensure that only tests performed during the infectious period contribute to a reduction in the infectious burden.

To cover the general case, where multiple tests may occur during the infectious period, only minor modifications to this description are needed.

The probability of testing positive at the *n*’th test, and (false) negative on all previous tests, is given by (1 − *s*)^*n*−1^*s*.

In order to compute the expected reduction *ρ*(*f, s*) due to the repeated testing scheme, we simply sum the contributions from the individual trajectories (being detected in the first test, the second test, and so on):

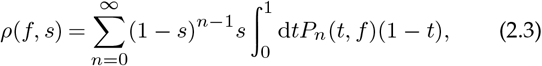

with *P*_*n*_(*t*; *f*) given by equation (2.1).

This expression can be computed analytically to yield

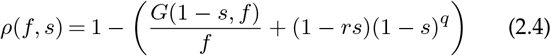

where the function *G*(*x, f*) is given by

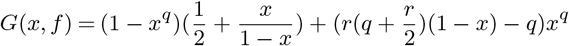

and *q* = ⌊*f* ⌋ is the integer part of the test frequency *f*, while *r* = *f* − ⌊*f* ⌋ = *f* mod 1 is the remainder.

### Incorporating heterogeneity

Heterogeneity in testing frequency *f* and social activity level *a* can readily be implemented in the mathematical framework. First, note that the burden of infection due to an individual with activity level *a* scales as *a*^2^ since activity modulates the risk of becoming infected (*r*_*S*_) as well as of transmission (*r*_*I*_), as introduced in the previous section. This can also be understood in terms of contact networks. Here, the reproductive number *R* is quadratic in connectivity; it depends on the product of in- and out-degrees. Since contacts are assumed symmetric – if individual *A* has an epidemiologically relevant contact with *B*, then *B* has one with *A* as well – the reproductive number of an individual with connectivity *c* may be written as

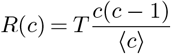

where *T* is the transmission risk per connection [28, 29]. In this description, connectivity is proportional to activity *a*, leading to an individual reproductive number which scales as *a*^2^.

The basic reproductive number *R*_0_ may then be computed as the average value

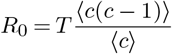

In the homogeneous mixing limit, where a very large number of connections are made and the transmission risk per connection is low, the above expression simplifies to

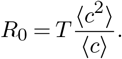

Given a joint distribution *P*_*b*_(*f, a*) of testing frequencies and activity levels in the population the expected reduction in the reproductive number is thus given by:

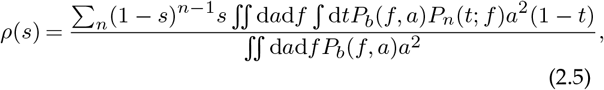

where *n* runs over 1, *…*, ∞, *t* runs over [0, 1] and the integrals over *a* and *f* both run over the entire real line.

#### Special Case: Testing and activity uncoupled

If testing behaviour and social activity levels are completely uncoupled, the distribution *P*_*b*_(*f, a*) factorizes, *P*_*b*_(*f, a*) = *P*_*f*_ (*f*)*P*_*a*_(*a*). In this case, the activity drops out of the expression for *ρ*(*s*) and the heterogeneity in activity thus has no bearing on the final result:

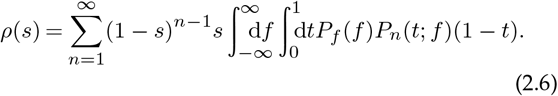

#### Special Case: Testing and activity perfectly coupled

The opposite extreme is the situation where activity and testing frequency are in direct proportion to one another, *a* ∝ *f*. The joint distribution can then be written as *P*_*b*_(*a, f*) = *P*_*f*_ (*f*)*δ*(*a* − *cf*) for some constant *c*. In that case, the expected reduction in the reproductive number is given by:

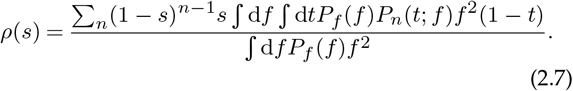

Note that the constant of proportionality *c* cancels and thus doesn’t affect the end result.

#### Parametrizing heterogeneit

In the limiting cases of independent or perfectly correlated test frequency and activity, we use the Gamma distribution *Γ* [*x*; *µ, k*] to describe the heterogeneity in either. Here *µ* is the mean value and *k* is the dispersion parameter which satisfies

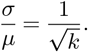

Here *σ* is the standard deviation of the distribution and *σ/µ* = *CV* is the coefficient of variation. The parameter *k* thus measures the homogeneity of the distribution, in the sense that *k* → ∞ corresponds to perfect homogeneity,

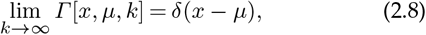

and low values, *k* < 1, correspond to high heterogeneity. For *k* = 1, the *Γ* distribution is simply an exponential distribution. For sufficiently small values of the dispersion factor, *k* approximates the metric “the most extreme fraction *f* of the population possesses 80% of the data” [30]. In other words, given an activity distribution with *k* = 0.1, one could say that “the most active 10% of the population account for 80% of the total activity”.

In the case of partially correlated frequency and activity, we generate the joint distribution by an algorithm which is described in the Supplemental Material. This algorithm ensures distributions of frequency of activity which have a controllable Pearson correlation coefficient as well as specified coefficients of variation and mean values.

## 3. Results and discussion

### Homogeneous populations – impact of test frequency and result delay

In a homogeneous testing scenario, the testing behaviour of the population is well-represented by a single number the average testing frequency. Likewise, we initially assume that the population is homogeneous with respect to social activity levels, and thus transmission rates. This regime is well-suited for exploring the impact of test-specific variables in isolation without the added complexity of a heterogeneous underlying population. We begin by addressing the impact of test result delay.

The time between testing and result availability varies by orders of magnitude between the types of tests commonly used for screening for SARS-CoV-2 – from a few minutes (e.g. rapid lateral flow antigen tests) to about a day (RT-PCR tests). In regular testing schemes, the tested individuals are generally not required to undergo isolation between test and result, and thus any delay will affect the total reduction of infection.

Delay can be taken into account directly, starting from the mathematical formulation presented in the Methods section. The only change required is to shift the time *t*_1_ of the first test by an amount *d*, i.e. letting *P*_1_(*t*_1_) → *P*_1_(*t*_1_ − *d*). Here, the delay *d* is measured in units of the infectious period *T*_*I*_. A slight reinterpretation of the variable *t*_1_ is also necessary it no longer strictly represents the time of the first test, but rather of the first test *result*, since it is this event that triggers isolation. The maximal reduction in reproductive number attainable with a test with a time delay is linear in the delay magnitude *d*, and the reduction *ρ*(*f, s, d*) in the delayed case is simply related to the instantaneous result:

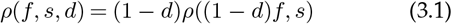

This relation reflects that the total number of test results obtained in the infectious period is diminished by a factor of (1 − *d*) (corresponding to letting *f* → (1 − *d*)*f*) while the expected reduction due to each of those test results is *also* reduced by the same factor (since they occur later in the infectious period), leading to the overall multiplication by (1 − *d*). As such, the dependence on delay duration is a nonlinear one.

In Figure 2, the reduction due to an instantaneous test is compared with a delayed one (at *d* = 0.2, corresponding to a one-day delay in a disease with a five-day infectious period). The reduction curves for the delayed tests thus tend toward a value of 80% as the frequency *f* is increased. For tests with a delay between test and result, it follows that arbitrarily large reductions cannot be obtained, and that increased test frequency cannot fully compensate for a delay.

**Figure 1.**
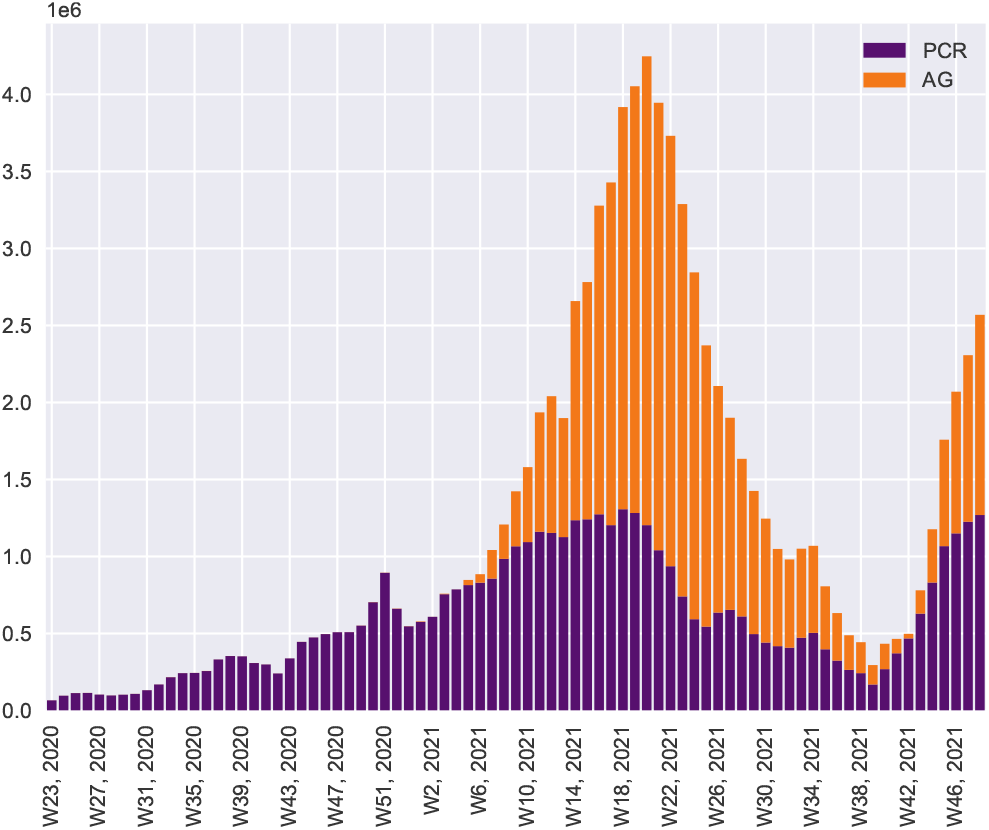
Overall level of testing over time, Danish data. Denmark has employed an extensive SARS-CoV-2 mass screening program. Assuming an infectious period of five days, the peak level of testing of approx. 4.4 million tests per week (attained around May 2021) corresponds to an average testing frequency of ⟨*f* ⟩ ≈ 0.5 in our model.

**Figure 2.**
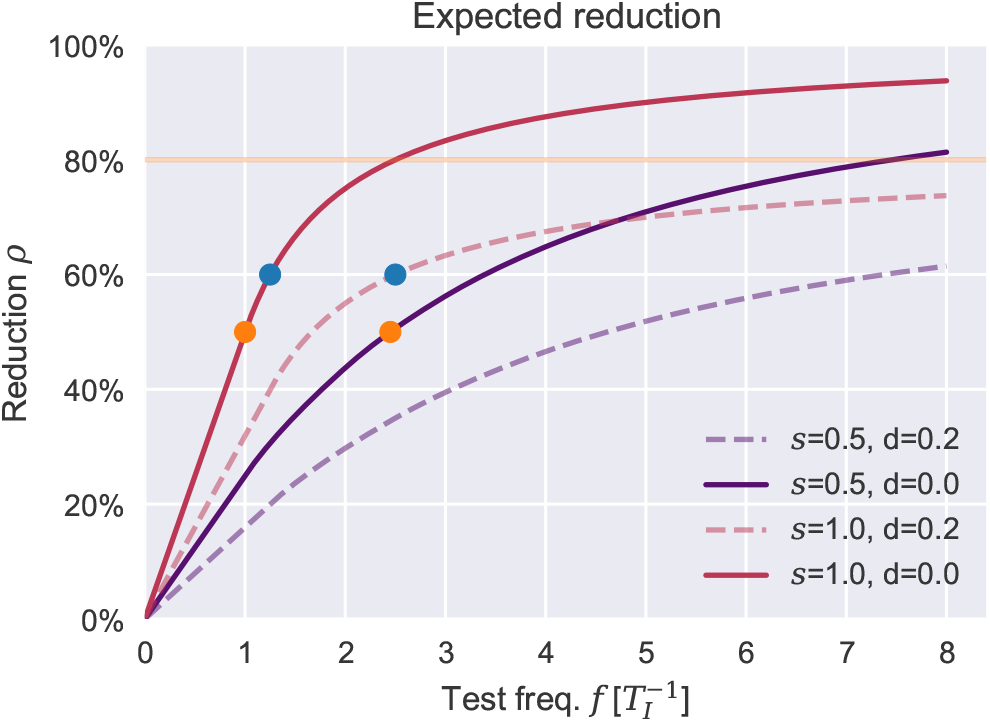
Efficient regular testing depends strongly on frequency and timeliness of results. The reduction in reproductive number obtained through regular testing depends strongly on the overall testing frequency. Concretely, assuming an infectious period of 5 days, a test with a sensitivity of just *s* = 50% performed at an interval of 2 days is as effective as a perfect-sensitivity test performed every 5 days (orange dots). However, even a high-frequency testing scheme suffers if results are delayed. Assuming a 5-day infectious period again, a delay of just one day (*d* = 0.2) has a sizable impact. Concretely, an instantaneous testing scheme performed every 4 days is as effective as delayed one performed every 2 days (blue dots). The dashed (delayed) lines asymptotically trend towards a maximum reduction value of 80% (orange horizontal line) with increasing test frequency. The delayed perfect-sensitivity test (dashed red line) only does so much more rapidly than its 50% sensitivity counterpart (dashed purple line)

As a function of testing frequency, the reduction *ρ* saturates at *ρ* = 1 − *d*, while it increases linearly for frequencies *f* < 1. Around *f* = 1*/*(1 − *d*), a law of diminishing returns kicks in, marking the point after which the reduction per test performed decreases. This is shown in Figure S2 of the Supplementary Material. As showcased by the initial linearity and eventual saturation of the reduction curves in Figure 2, the effectiveness of different test scenarios generally depend on the parameters in a highly nonlinear fashion. In the figure, two pairs of dots mark pairs of equally effective testing scenarios which differ in test sensitivity and delay-to-result time, respectively. The aforementioned non-linearity is clear when considering the growing distance between each such pair of curves as the reduction level varies.

### Heterogeneity in testing impedes mitigation but reduces the importance of high test sensitivity

Populations are rarely homogeneous where behaviour is concerned, and rates of testing as well as contact rates are likely to vary widely [31]. In this section, we explore the impact of heterogeneous testing behaviours on its own - that is, without any correlation to social activity. As described in the Methods section, any variability in contact rates are immaterial in this case, and can be ignored for now.

In order to directly gauge the impact of heterogeneity, we parametrize the testing frequency by a Gamma distribution with a controllable dispersion factor *k* and a mean value ⟨*f* ⟩, as described in the Methods section. A lower *k* value corresponds to a highly heterogeneous distribution. *k* = 1 can be viewed as a cross-over value between the highly heterogeneous regime (*k* < 1, where the spread is larger than the mean value) to the fairly homogeneous case (*k* > 1, where fluctuations are typically smaller than the mean).

As shown in Figure 3a, an increase in dispersion (smaller *k*) leads to a drop in effectiveness of mitigation, measured as the reduction in reproductive number, even if the total number of administered tests is similar. In other words, regular testing as a mitigation strategy becomes less cost-effective in the face of heterogeneous testing behaviour. This overall result can be understood in the following way. First, in a heterogeneous scenario, a large proportion of the population are tested very rarely. Secondly – and more importantly – at the other extreme are a group who are tested so frequently that each additional test only contributes relatively little to the expected probability – and time – of detection. This effect is visible for both cases of sensitivity, *s* = 0.5 and *s* = 1.0, but most pronounced for the ideal test, *s* = 1. (Figure 3a, red curves).

**Figure 3.**
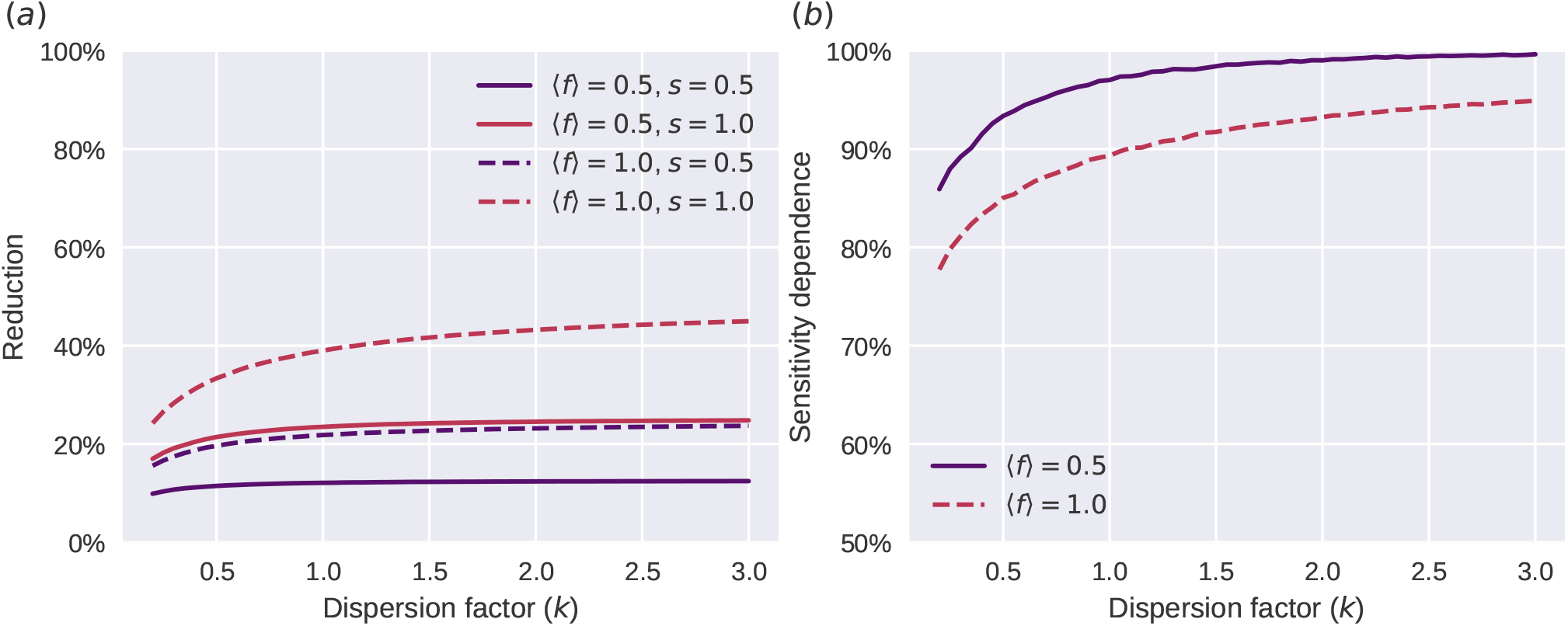
Heterogeneous testing behaviours impede mitigation when testing frequency is heterogeneous and uncorrelated with social activity. Note that test sensitivity becomes less important as heterogeneity increases. **(a)**: Reduction as function of dispersion coefficient. The fully drawn lines are for *f* = 0.5 (purple), the dashed lines are *f* = 1.0 (red), and the two colors are for resp. *s* = 0.5 and *s* = 1.0 **(b)**: The dependence of the reduction due to the test-sensitivity decreases as heterogeneity increases (as *k* → 0)

In Figure 3b, we explore the role that test sensitivity plays in modulating the overall mitigative power. Naturally, test sensitivity is an important parameter in shaping the efficacy of a test programme, but a less-than-ideal sensitivity can be largely offset by an increased frequency of testing (see Figure 2 and ref. [17]). The intuition behind this phenomenon is that the probability to remain undetected throughout the infectious period is reduced exponentially as a function of the number of tests performed. Assuming a test-sensitivity of *s*, the probability *p*_*n*_ that a positive individual has been detected after *n* tests is then

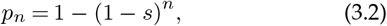

assuming that each test is an independent event. However, this simple description holds only on the scale of a single individual. If a fixed number of tests are instead heterogeneously distributed in a population, it is not *a priori* obvious how much of a role the sensitivity plays.

We quantify the sensitivity dependence using the following measure:

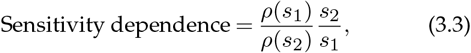

where *s*_1_ and *s*_2_ are two different sensitivities and *ρ* is the expected reduction in reproductive number. A sensitivity dependence of 100% indicates that decreasing test sensitivity by some factor leads to a decrease in mitigation by that same factor. A sensitivity dependence of < 100% thus indicates a reduced vulnerability to less-than-ideal test sensitivity. The curves of Figure 3b arise by comparing the reduction obtained at a sensitivity of *s*_1_ = 100% to that obtained at a sensitivity of *s*_2_ = 50% by means of the above equation. Clearly, test sensitivity plays less of a role when testing is heterogeneous (low *k*). As we shall see in the next section, this result survives – and is in fact strengthened – when heterogeneous testing behaviour is correlated with social activity.

Intuitively, the result can be explained in the following way. In a heterogeneous testing scenario, a large proportion of the population very rarely get tested, and so are not strongly affected by a decrease in test sensitivity. In the case of a dispersion of e.g. *k* = 0.2, the majority of tests are taken by the upper 20% of the population who get tested so often that, again, test sensitivity is not a grave concern since it is offset by the high frequency. In a homogeneous scenario, on the other hand, the majority of individuals are tested at an intermediate rate, where each false negative is likely to have a real impact on how early the pathogen is detected – if detected at all.

The decreased efficiency of regular testing schemes in the face of heterogeneous testing behaviours is exacerbated by the fact that those individuals who only get tested rarely do not necessarily have a correspondingly low risk of infection or transmission. At the opposite extreme, the section of the population who get tested frequently are not guaranteed to be the most socially active and are therefore not expected to account for the majority of new infections anyway. In other words, the lack of correlation between test frequency and social activity (with its associated exposure risk), is exactly what makes a heterogeneous testing scheme perform so relatively poorly.

In the next section, we explore how a regular testing scheme performs when test frequency and social activity *are* correlated.

### Test/activity correlation renders heterogeneity an advantage

In the previous section, we assumed that testing rates were heterogeneously distributed, but completely uncorrelated with social behaviour in general. We now turn to the other extreme and assume that the social activity (or contact rate) is fully correlated with the test-frequency – that they are directly proportional. The plots of Figure 4 were generated by evaluating equation (2.7) under this assumption. Clearly, introducing such a correlation radically alters the effects of heterogeneity on the mitigation strategy. By inducing correlation, heterogeneity can in fact be leveraged to significantly improve the performance of a regular testing scheme.

**Figure 4.**
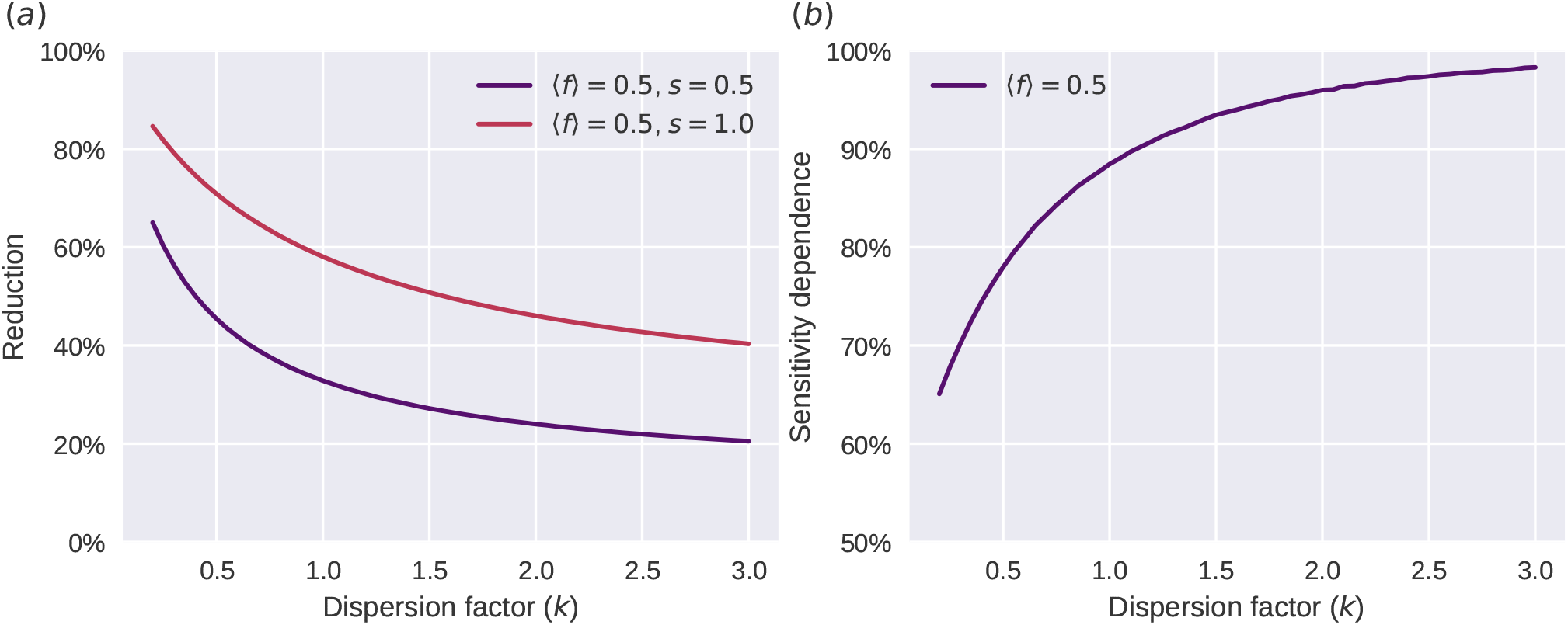
Testing heterogeneity can be turned to an advantage. Here, perfect correlation between social activity and test frequency is assumed. **(a)** (left): Reduction in the reproductive number as a function of the dispersion coefficient *k* (lower *k* corresponding to more pronounced heterogeneity). Note that for the fully correlated case, increased heterogeneity leads to better epidemic control (higher reduction in the reproductive number), in contrast to what was observed in the uncorrelated case (Figure 3). **(b)** (right): The reduction due to regular testing depends less strongly on test sensitivity when heterogeneity is high, in the fully correlated case. The overall dependence is similar to the one observed for the uncorrelated case (Figure 3), only more pronounced.

Furthermore, the trend observed in the uncorrelated case with respect to test sensitivity continues to hold here. The more heterogeneous testing and activity becomes, the less discrepancy between the performance of high- and low-sensitivity tests is observed.

Of course, neither of these extremes are likely to exactly represent any real scenario, but a better description probably exists somewhere in between, with an incomplete but nonzero correlation between social and testing activity. We generate partially correlated distributions of activity and test frequency with a specified level of dispersion using an algorithm which is described in the supplementary material. We continue to express the level of dispersion in terms of the parameter *k* = (*µ/σ*)^2^. Once the distributions have been generated, the reduction due to testing can be computed using equation (2.5). This procedure results in Figure 5, which systematically explores the relation between the degree of test/activity correlation (as measured by the Pearson correlation coefficient) and the expected reduction in reproductive number. We find that even a very weak correlation renders heterogeneity an advantage for a regular testing scheme, and that the effect is rather dramatic even at moderate correlation levels.

**Figure 5.**
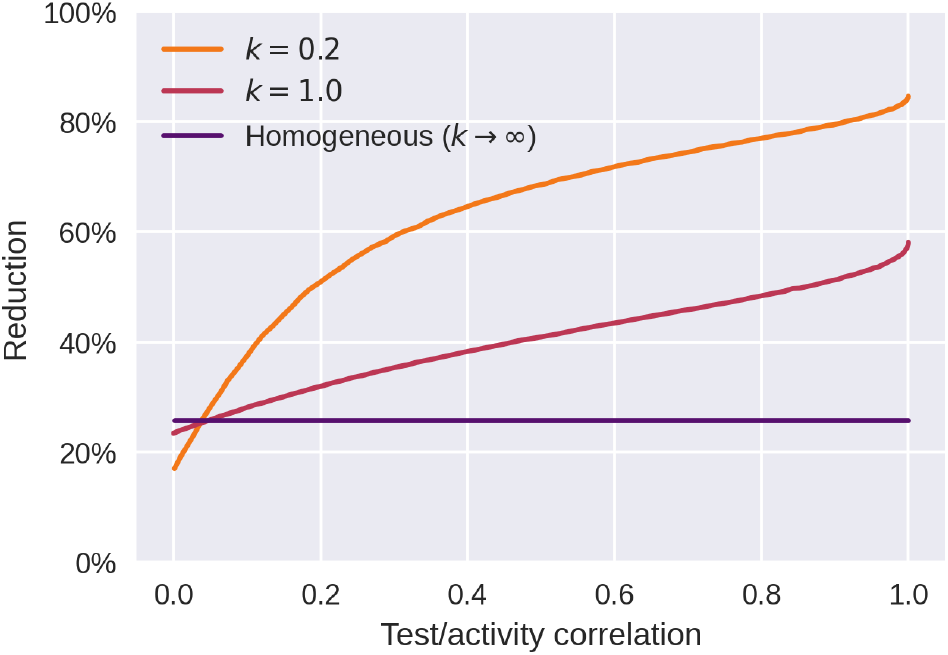
Only weak coupling is needed to leverage heterogeneity. Dependency of reduction on the correlation between test frequency and activity. Across all three curves, the test frequency is set at *f* = 0.5, and the test sensitivity is assumed ideal, i.e. *s* = 1. Note that the turning point at which heterogeneity becomes an advantage rather than a disadvantage occurs at quite low correlations – as long as a correlation of more than ≈ 5% is attained, the heterogeneous scenarios (*k* = 0.2 and *k* = 1.0) fare better than the homogeneous one (*k* → ∞).

### Perspectives

In addition to screening and direct isolation, testing schemes play a role in enabling contact tracing for close contacts to infectious individuals. An oft-used abbreviaton is *TTI* – Test, Trace, Isolate. Here we have only addressed the effects of the screening aspect and subsequent isolation, but not any kind of contact tracing schemes which may be implemented in addition. As addressed in refs. [30, 32, 33], overdispersion in infectiousness can be leveraged to improve contact tracing, by incorporating a bidirectional (backward-then-forward) contact tracing scheme. This effect relies on an analogy to the so-called friendship paradox of network theory [34]. This is the somewhat counter-intuitive statement that “on average, your friends have more friends than you do”, which holds true as long as the underlying degree distribution has a nonzero variance. Similarly, if reproductive numbers in a disease outbreak are variable, then it is true that “the person who infected you likely infected more people than you did”. However, even simple forward contact tracing benefits from heterogeneities in social activity and contact network structure as well [35].

If activity is *correlated* with test frequency, as described in this study, contact tracing is likely to receive a significant boost. In this case, the heterogeneous activity leads to a friendship paradox, as described above, while the coupling to test frequency increases the likelihood of *detecting* primary cases with a high risk of being infected in the first place. We thus highlight the need for research to establish the exact impact of (correlated) heterogeneous activity and test frequency on contact tracing schemes. Such research is likely to yield insights into how test-based COVID Passport-type solutions can increase the effectiveness of contact tracing as well as decrease the overhead associated with this type of control strategy.

The efficacy of contact tracing schemes depends on the characteristics of the test employed as well. In this context, high test specificity is mainly a question of minimizing the overhead caused by false positives and of staying within the capacity of the infrastructure surrounding the contact tracing scheme. Since contact tracing is a time-sensitive operation, any delay from test to result, as well as in tracing itself, has a large impact on the efficacy of contact tracing [36, 37].

In this study, we have assumed that all heterogeneity in susceptibility and infectiousness stems from differences in social activity. Put differently, we have not taken potential biological heterogeneities into account. However, it is well-known that certain infectious diseases are prone to high person-to-person variability in infectiousness, with COVID-19 being an example of such a disease [38–44]. It is also becoming increasingly clear that biological variability plays a significant role in explaining this disease-specific overdispersion in infectiousness [45–48] and that the resulting *superspreading* phenomenon has wide implications for mitigation strategies [30, 32, 38, 49–53]. A central finding has been that overdispersion in infectiousness enhances the sensitivity of an epidemic to changes in social network size and structure [30, 50], a link that has yet to be explored in the context of regular testing schemes.

### Limitations

While our model improves upon previous mathematical models of regular testing programmes by including heterogeneities and correlations, it makes a number of idealizations. The model assumes that the onset of the infectious period coincides with the earliest time at which the pathogen is detectable by the screening test employed. This is only a coarse-grained approximation, the validity of which depends on the exact test and pathogen in question. In the case of SARS-Cov-2, studies into the connections between viral load (a proxy for infectiousness) and probability of detection suggest that detectability precedes infectiousness, at least for high-sensitivity tests [17, 54, 55]. For lower-sensitivity tests such as lateral flow antigen tests, the approximation is likely to be more accurate. In any case, early detectability would of course be a benefit to a regular testing scheme. Furthermore, high-sensitivity RT-PCR tests are more likely than antigen tests to detect fragments of non-viable virus for a prolonged period after the individual is no longer infectious [56]. These false positives of course contribute nothing in terms of mitigation but have no bearing on our results.

It should be noted that this study does not consider the wider range of interventions (pharmaceutical and otherwise) which are likely to affect testing frequency. In the case of SARS-CoV-2, the most notable example is the vaccine roll-out. Testing requirements associated with COVID Passport solutions are often affected by vaccination status [24, 25], leading to correlations between vaccination status, rate of infection and frequency of testing.

We have generally assumed perfect regularity in the spacing between tests. However, in the Supplementary Material, we explore the opposite extreme: stochastic timing of tests with no dependence on the time of the previous test, corresponding to a Poisson process model of testing (see Figure S1 of the Supplementary Material). We find that the qualitative agreement is good, and thus expect our results to have quite general applicability, irrespective of the precise details of the underlying testing scheme.

Finally, regular testing is a type of screening and thus only targeted at symptom-free individuals. As such, the population-wide impact of regular testing is highly dependent on the presymptomatic period as well as the asymptomatic fraction for the disease in question. In this study, we model only the mitigation impact among subpopulations who *do* participate in regular testing, and not the society-wide impact of such a testing programme.

## 4. Conclusions

Person-to-person heterogeneity is increasingly recognized as a decisive factor in many epidemiological phenomena. We have shown that heterogeneous testing behaviour, in and of itself, is disadvantageous to regular testing schemes. This result largely owes to a basic property of overdispersed distributions; when significant heterogeneity is present, a large fraction of the population is essentially non-participatory while some individuals undergo very frequent testing. When a fraction of the population is tested very frequently, each additional test contributes less in terms of epidemic control than if it were redistributed to less-frequently tested individuals.

Heterogeneity was also shown to alter the population-wide impact of properties inherent to the test itself. Our results show that the *sensitivity* of the test involved is less important in the case of heterogeneous testing. It was previously shown by Larremore et al. [17] that even tests with a moderate sensitivity are highly useful in (homogeneous) regular testing schemes, provided that sufficiently rapid result availability and testing frequency is possible. Our results thus strengthen this finding by showing that heterogeneity further increases the effect. This decreased dependence on test sensitivity is a robust finding and continues to hold in the case of correlated testing and activity distributions discussed below.

As long as heterogeneity in testing and in general social activity are uncoupled, the effect of testing heterogeneity is a detrimental one. However, correlations between activity and testing are natural to consider and are even induced through the design of certain COVID Passport solutions where a recent negative test is required to hold a valid COVID Passport. Such a passport in turn grants entry to many aspects of public life, ranging from educational institutions and public transport to bars and restaurants. We have shown that heterogeneous testing behaviours can in fact be leveraged to increase the mitigation effect of a population-wide regular testing programme. By coupling testing frequency to social activity, heterogeneity turns from a disadvantage to a significant advantage. Concretely, just a weak correlation (< 10%) between activity and testing is necessary to benefit from heterogeneity, and the effect increases rapidly with enhanced correlation. Our work thus provides a theoretical basis for the design of test-**9** based “passport” solutions. Crucially, the low threshold correlation required to reap the benefits of heterogeneity indicate that even initiatives which moderately couple testing to activity may be highly beneficial.

## Data Availability

All data and computational code produced in the present study are available upon reasonable request to the authors.

## Data Accessibility

Insert data access text here.

## Acknowledgements

We thank Lone Simonsen, Andreas Eilersen and Kim Sneppen for enlightening discussions.

