## Supplementary Material for "Heterogeneity in testing for infectious diseases"

### Heterogeneity in testing for infectious diseases: Supplementary material

(Dated: January 11, 2022)

#### I. GENERATING PARTIALLY CORRELATED DISTRIBUTIONS

In this section, we describe a numerical procedure for generating two correlated distributions with specified mean values $\mu_i$ and dispersion factors $k_i$ (understood in a generalized sense as the square of the reciprocal coefficient of variation, $k = (\mu/\sigma)^2$, with $\mu$ the mean value and $\sigma$ the standard deviation).

We shall refer to the two stochastic variables as activity $a$ and frequency $f$, since they represent social activity levels and testing frequencies in the main text. The population size is taken to be $N$, and as such a distribution of activities is realized as a vector $\mathbf{a} = (a_1, \ldots, a_N)$, and likewise for the frequencies, $\mathbf{f} = (f_1, \ldots, f_N)$.

The first part of the algorithm consists of drawing initial (uncorrelated) frequencies $\mathbf{f}$ and activities $\mathbf{a}$, and then creating a third distribution $\tilde{\mathbf{f}}$ which has a partial correlation with $\mathbf{a}$ (the magnitude of which is controlled by an auxiliary parameter $\xi$):

- Draw a vector $\mathbf{f}$ from a Gamma distribution $\Gamma[f; \mu_f, k_f]$
- Draw a vector $\mathbf{a}$ from a Gamma distribution $\Gamma[a; \mu_a, k_a]$
- Let $\tilde{\mathbf{f}} = (1-\xi)\mathbf{f} + \xi\mathbf{a}$, with $\xi \in [0; 1]$.

The next part of the algorithm is iterative and proceeds as follows. Note that $CV(\mathbf{x}) = \langle \mathbf{x} \rangle / \sqrt{\mathrm{Var}[\mathbf{x}]}$ is a function which computes the coefficient of variation of the elements of a vector (or tuple) $\mathbf{x}$.

- Let the desired coefficient of variation be $\chi = 1/\sqrt{k_f}$ and the tolerance be $\mathrm{d}\chi$.
- While $|CV(\tilde{\mathbf{f}}) - \chi| > \mathrm{d}\chi$:
  - Let $z = \mathrm{sgn}(CV(\tilde{\mathbf{f}}) - \chi)$
  - Let $\tilde{\mathbf{f}} \to \tilde{\mathbf{f}} - z\varepsilon(\tilde{\mathbf{f}} - \langle\tilde{\mathbf{f}}\rangle\mathbf{1})$ to increase or reduce the spread of the distribution, depending on the value of $z = \pm 1$. Here, $\mathbf{1}$ denotes the vector $(1, 1, \ldots, 1)$ with $N$ elements.
- Let $\tilde{\mathbf{f}} \to (\mu_f/\langle\tilde{\mathbf{f}}\rangle)\tilde{\mathbf{f}}$, to rescale $\tilde{\mathbf{f}}$ to have the desired mean value.

Here, $\varepsilon$ is a small quantity which sets the rate of convergence. The tolerance $\mathrm{d}\chi$ is the maximal acceptable deviation from the desired coefficient of variation. This procedure results in a distribution $\tilde{\mathbf{f}}$ which has a mean value $\mu_f$, a dispersion factor $k_f$ and a partial correlation with $\mathbf{a}$. As the parameter $\xi$ varies between 0 and 1, the Pearson correlation coefficient between $\tilde{\mathbf{f}}$ and $\mathbf{a}$ likewise varies between 0 and 1. The exact relation between $\xi$ and the correlation $\rho$ can be derived as follows. The joint distribution of $f$ and $a$ is simply

$$P_{af}(a, f) = P_a(a)P_f(f), \tag{1}$$

since $a$ and $f$ are independent. To find instead the joint distribution of $a$ and $\tilde{f}$, we express $f$ in terms of $a$ and $\tilde{f}$:

$$f(\tilde{f}, a) = \frac{\tilde{f} - \xi a}{1-\xi}. \tag{2}$$

Recalling that $P_{af}(a, f)$ is a probability *density*, the integration measure must also change. That is (using the above relation for $f(\tilde{f}, a)$),

$$P_{af}(a, f(\tilde{f}, a))\mathrm{d}a\mathrm{d}f = P_{af}\left(a, \frac{\tilde{f} - \xi a}{1-\xi}\right)\frac{\mathrm{d}a\mathrm{d}\tilde{f}}{1-\xi} \tag{3}$$

$$\equiv P_{a\tilde{f}}(a, \tilde{f})\mathrm{d}a\mathrm{d}\tilde{f}. \tag{4}$$

In other words, the joint distribution of $a$ and $\tilde{f}$ is:

$$P_{a\tilde{f}}(a, \tilde{f}) = \frac{1}{1-\xi}P_a(a)P_f\left(\frac{\tilde{f} - \xi a}{1-\xi}\right) \tag{5}$$

The correlation between $a$ and $\tilde{f}$ can then directly be computed as:

$$\begin{aligned}\rho &= \frac{\mathrm{Cov}[a, \tilde{f}]}{\sqrt{\mathrm{Var}[a]\mathrm{Var}[\tilde{f}]}} \\ &= \frac{\iint (a-\mu_a)(\tilde{f} - \mu_{\tilde{f}})P_{a\tilde{f}}(a, \tilde{f})}{\sqrt{\iint (a-\mu_a)^2 P_{a\tilde{f}}(a, \tilde{f})\mathrm{d}a\mathrm{d}\tilde{f} \iint (\tilde{f} - \mu_{\tilde{f}})^2 P_{a\tilde{f}}(a, \tilde{f})\mathrm{d}a\mathrm{d}\tilde{f}}} \\ &= \frac{\xi}{\sqrt{1 + 2(\xi - 1)\xi}}\end{aligned} \tag{6}$$

This relation can be inverted to yield $\xi$ as a function of the desired correlation $\rho$:

$$\xi(\rho) = \frac{\rho^2 - \sqrt{\rho^2 - \rho^4}}{2\rho^2 - 1}. \tag{7}$$

*
†
‡

Using this relation, and the procedure just described, partially correlated distributions of activity and test frequency can be generated at any level of correlation desired.

#### II. A POISSON PROCESS MODEL OF TESTING

In the main text, we have assumed that testing occurs at perfectly regular intervals, such that the time of each test follows deterministically from the previous one. An alternative assumption is that there is, at each instant of time, a constant probability rate for undergoing testing. In such a constant-rate scheme, testing is described by a Poisson process, which in turn leads to an exponentially distributed waiting time. In this section, we explore such a model of heterogeneous testing, in which the underlying testing rate of each individual is given by the frequency $f$.

We first derive a few analytical results for the cases of (i) heterogeneous testing (no correlation) and (ii) heterogeneous testing and activity, perfectly correlated. We then show, by means of computer simulation, that the constant-rate model qualitatively agrees with the perfectly regular testing model used in the main text. The constant-rate approximation is often a convenient assumption in computer models, since it has the property of being memory-free; the probability of testing is independent of the time that has elapsed since the last test was performed.

##### A. Heterogeneous testing, homogeneous activity

We assume a test with perfect sensitivity and furthermore that

- The testing rate $f$ in the population is distributed according to a (Gamma) distribution $P_f[f;k,\mu]$
- As described above, testing itself is assumed to be a constant-rate process. It follows that the time of testing is then distributed according to the exponential distribution $P_t[t,f]$, with $f$ the rate constant (as drawn above).

As in the main text, it is assumed that the level of infectiousness is constant throughout the infectious period. For a single individual $i$ who is tested at time $t_i$ during their infectious period (of unit total duration), the prevented fraction of infections is then $\Delta S_i = 1 - t_i$. The average prevented fraction of infections is thus given by:

$$\langle \Delta S \rangle = \int_{f=0}^{\infty} \int_{t=0}^{1} P_f[f;k,\mu] P_t[t;f](1-t) \, \mathrm{d}f \, \mathrm{d}t \tag{8}$$

When $P_f$ is taken to be a Gamma distribution with dispersion parameter $k$ and mean $\mu$, the above expression can be evaluated in closed form to yield:

$$\langle \Delta S \rangle = \frac{k(\mu-1) + k^k (k+\mu)^{1-k} - \mu}{\mu(k-1)} \tag{9}$$

While this expression has a singularity at $k=1$, it is removable, and the limit is well defined:

$$\lim_{k\to 1} \langle \Delta S \rangle = 1 - \frac{1}{\mu} \log(1+\mu) \tag{10}$$

##### B. Heterogeneous and (perfectly) correlated test + activity

Here, we assume that the activity levels (contact rates) $a$ of individuals are heterogeneous and given by a Gamma distribution $P_a[a;k,\mu]$. Since the contact rate determines the risk of becoming infected as well as the risk of passing on infection, the infectious potential of an individual is proportional to $a^2$.

Under these assumptions, the mean prevented infectious load can again be computed analytically:

$$\begin{aligned}
\langle \Delta S \rangle &= \int_{a=0}^{\infty} \int_{t=0}^{1} P_a[a;k,\mu] P_t[t;f(a)] a^2 (1-t) \, \mathrm{d}a \, \mathrm{d}t \times \left[ \int_{a=0}^{\infty} P_a[a;k,\mu] \; a^2 \; \mathrm{d}a \right]^{-1} \\
&= \frac{1}{\Gamma[2+k]} e^{-f_{\max}} k \mu^{-2-k} (k+\mu)^{-1-k} \{ (k+\mu)(f_{\max} k(k+\mu))^k \left( \mu + k\mu + e^{f_{\max}} (f_{\max} k - (1+k)\mu) \right) E_{-k}[f_{\max} k/\mu] \\
&+ \mu \Big( - e^{-f_{\max} k/\mu} (e^{f_{\max}} - 1)(k+\mu)(f_{\max} k(k+\mu))^k + e^{f_{\max}} \mu^k \big( (k+\mu)^{1+k} (k(\mu-1)+\mu) \Gamma[k] \\
&+ k^{1+k} (\Gamma[1+k] - \Gamma[1+k, (f_{\max}(k+\mu))/\mu]) \big) \Big) \}
\end{aligned} \tag{11}$$

where $\Gamma[a,z]$ is the incomplete Gamma function:

$$\Gamma[a,z] = \int_z^{\infty} t^{a-1} e^{-t} \mathrm{d}t$$

and $E_n[z]$ is the exponential integral function:

$$E_n[z] = \int_1^{\infty} e^{-zt}/t^n \mathrm{d}t. \tag{12}$$

In Figure S1, we show the results of numerical simulations of a Poisson process model for testing with heterogeneity, echoing the plots of Figures 3a and 4a in the main text. The model is implemented in an *agent based* fashion, in the sense that each individual is equipped with an activity level and test rate $f$ and then participates in a regular testing scheme. However, no interaction *between* agents is modeled, and thus the model is extremely simple. The activity and frequency distributions are generated using the procedure described in the previous section, and may thus be either uncorrelated (panel a), fully correlated (panel b), or even partially correlated. The trends observed in Figure S1 are very similar to those obtained in the main test with a fixed-interval testing model. Our results regarding the impact of heterogeneity and correlation are thus quite robust and not tied to any particular model of the underlying testing scheme.

#### III. ANALYTICAL TREATMENT OF HOMOGENOUS TESTING

In this section, we present an alternative derivation of the basic equations of regular testing. In the main text, we developed a framework in which we made use of Dirac's delta function to compute the expected reduction in reproductive number. Here we present an alternative derivation which, while not as brief, may be more intuitive.

##### A. The regular screening

The total duration of infectiousness is $T_I$. In order to compute the expected number of tests performed during the infectious period, we define the following auxiliary quantities:

$$f = \frac{T_I}{T_R} \quad , \quad q = \lfloor f \rfloor \quad , \quad r = f \mod 1$$

where $\lfloor x \rfloor$ denotes the floor function and $d < 1$ the result delay in units of $T_I$. The quantity $f$ is thus frequency of testing in units of the $T_I^{-1}$. $f$ can be decomposed into its integer part $q$ and remainder $r$ as $f = q + r$.

The number of tests being performed during an infectious period can then be expressed in terms of the quantities defined above. The probability of being tested with $q$ tests over the infectious period $T_I$ is $(1 - r)$ and the probability of getting $q + 1$ tests is $r$.
Let $m$ denote the number of tests being performed during the infectious period. This number implicitly depends on $f$. The maximum number of tests possible during the infectious period, can be expressed as:

$$P(m = q) = (1 - r)$$
$$P(m = q + 1) = r$$

The expected number of tests during the infections period is thus given by

$$\langle m \rangle = rq + (1 - r)(q + 1) = 1 + q - r$$

since only the last two terms contribute to this sum.

##### B. Probability of detection

One can now calculate the probability of being tested positive in the infectious period, given that a person is actually infectious. This requires the additional insight that a person is tested positive on either first test, 2. test, etc. until the last test ($q$ or $(q + 1)$) is performed.

Let $D$ denote the outcome, that the infected person is detected and $\neg D$ the negation. $I$ (for infectious) denotes the true state of the individual is infectious and its negation $\neg I$

Introducing the sensitivity of the test $s$, then:

$$s = P(D|I, n = 1)$$
$$(1 - s) = P(\neg D|I, n = 1)$$

where $n$ here is introduced as the test number; the $n = 1$ in the expression above means that we only consider a single test event. These probabilities corresponds to respectively the true positive and false negative rates of the test.

Now the probability of being detected during the entire infectious period an be calculated, from considering the negation;

$$P(\neg D|I) = 1 - P(D|I)$$

With probability $(1 - r)$, a maximum of $q$ tests are performed and with probability $r$, a maximum of $q + 1$ tests are preformed. Negating the probability of no detection, results in:

$$P(D|I) = 1 - \left((1 - r)(1 - s)^q + r(1 - s)^{q+1}\right)$$
$$= 1 - (1 - rs)(1 - s)^q$$

##### C. Time of detection

The first test, $t_0$, is assumed distributed in time as a uniform distribution $t_0 \sim \frac{1}{T_R}$ and will therefore have an average value of $\frac{T_R}{2}$. Thus the (average) time at which the $n$'th test is performed, $t_n = nT_R + t_0$, must therefore be:

$$\frac{1}{T_R} \int_0^{T_R} (t_n) \mathrm{d}t_0 = (n + \tfrac{1}{2}) T_R \qquad (13)$$

for $n \leq q$. Special care need to be taken in regard to the (potentially) last test (test $q + 1$). Since the last test will

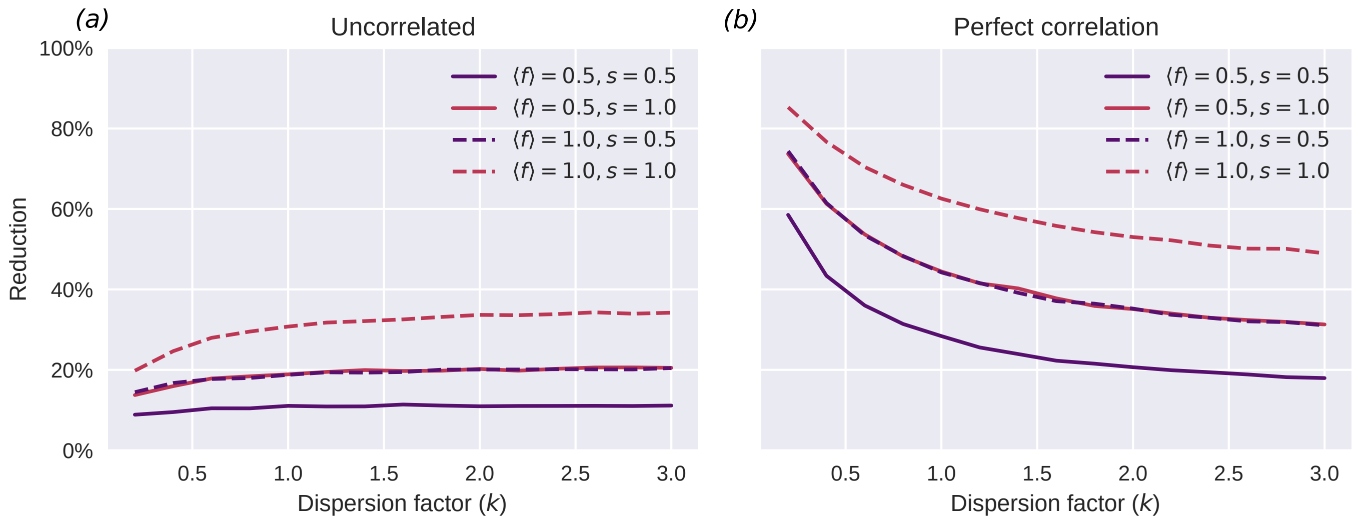


FIG. S1. A Poisson (constant-rate of testing) model of heterogeneous, regular testing performs qualitatively similarly to the fixed-interval model of the main text. **a)** Uncorrelated test frequency and activity distributions **b)** Perfectly correlated test frequency and activity distributions.

be taken at time $t_q = qT_R + t_0$ where this time $t_0 \sim \frac{1}{rT_R}$ uniformly distributed, we have that:

$$\frac{1}{rT_R}\int_0^{rT_R}(t_q)\mathrm{d}t_0 = \frac{1}{rT_R}\int_0^{rT_R}(qT_R + t_0)\mathrm{d}t_0 = \left(q + \frac{r}{2}\right)T_R$$

The expectation value of time before detection, $t_D$, when a detection is actually occurring, is thus:

$$\mathbb{E}[t_D, D|I](f,s) = \left[\theta(q-\frac{1}{2})\sum_{m=0}^{q-1}\left((m+\frac{1}{2})(1-s)^m\right) + r(q+\frac{r}{2})(1-s)^q\right]sT_R$$

The expression can be simplified into a closed form by considering the following function (calculated via methods from generating functions):

$$F(x,f) = \sum_{m=0}^{q-1}(\frac{1}{2}+k)x^m = \frac{1-x^q}{1-x}\left(\frac{1}{2}+\frac{x}{1-x}\right) - q\frac{x^q}{1-x}$$

Identifying $x = 1-s$, we get:

$$F(1-s,f) = \frac{1-(1-s)^q}{s}\left(\frac{1}{2}+\frac{1-s}{s}\right) - q\frac{(1-s)^q}{s}$$

Since there is now no ambiguity for $q = 0$, the expectation value can now be written as:

$$\begin{aligned}\mathbb{E}[t_D, D|I] &= \left(F(1-s,f) + r(q+\frac{r}{2})(1-s)^q\right)sT_R \\ &= G(1-s,f)T_R\end{aligned}$$

where

$$G(x,f) = (1-x^q)(\frac{1}{2}+\frac{x}{1-x}) + (r(q+\frac{r}{2})(1-x) - q)x^q$$

##### D. Reduction of infection

Since the time of detection is a measure for the spread of infection, the reduction of infectiousness compared with no intervention for asymptomatic individuals can be calculated as:

$$\rho(f,s) = \frac{T_I - \mathbb{E}[t_D|I](f,s)}{T_I} = 1 - \frac{\mathbb{E}[t_D|I](f,s)}{T_I} \quad (14)$$

where $t_D$ is the time of detection after onset of infectiousness. The extra arguments $(f,s)$ in the expectation value above, is just to make the dependency of detection time on sensitivity and frequency clear. The expression that give the reduction is thus:

$$\begin{aligned}\mathbb{E}[t_D|I] &= \mathbb{E}[t_D, D|I] + \mathbb{E}[t_D, \neg D|I] \\ &= \mathbb{E}[t_D, D|I] + \mathbb{E}[t_D|\neg D, I]P(\neg D|I) \\ &= \mathbb{E}[t_D, D|I] + T_I P(\neg D|I)\end{aligned}$$

due to $T_I$ being the time attributed to undetected individuals w.r.t. a reduction perspective; the maximal time one can infect. Using the relation found earlier:

$$\frac{\mathbb{E}[t_D|I](f,s)}{T_I} = G(1-s,f)\frac{T_R}{T_I} + P(\neg D|I)$$

whereby the reduction is now found as:

$$\rho(f,s) = 1 - \left(G(1-s,f)\frac{1}{f} + (1-rs)(1-s)^q\right)$$

##### E. Impact of test frequency and result delay

Using the same argument as in the main text or considering a scaling of the frequency to accommodate the delay of the test-result yields the same expression. Here We give the second argument. when the results are delayed the effective frequency $f_d$ is scaled such that the frequency in relation to the reduced period in which one can receive the test-results (and thus play the same game as for the no-delay test):

$$f_d = \frac{T_I - dT_I}{T_R} = (1-d)f$$

Furthermore, a fraction $d$ of the infectious period is reserved for result delay (one cannot receive a result before the time $dT_I$), we therefore find that, when taking into account the result-delay, the reduction becomes:

$$\begin{aligned}
\rho(f,s,d) &= 1 - \left(\frac{\mathbb{E}[t_D|I](f_d,s) + T_I d}{T_I}\right) \\
&= (1-d) - \frac{\mathbb{E}[t_D|I](f_d,s)}{T_I} \\
&= (1-d)\left(1 - \frac{\mathbb{E}[t_D|I](f_d,s)}{T_I(1-d)}\right) \\
&= (1-d)\rho((1-d)f,s)
\end{aligned}$$

Applying the analytical result found for $\rho(f)$ and finding (numerically) the derived function (wrt. $f$), we can gain a visualization of the measure "reduction-per-test" from the resulting figure S2 as seen below:

From these curves, we see that the "diminishing return effect" starts to have effect only after the condition $f(1-d) = 1$ is met, since this is the effective frequency at which a single individual is ensured to receive the result of the first test, within the infectious period. Thus the maximal reduction possible under the constraint that we should also maximize the "reduction-per-test"-measure happens at $f = \frac{1}{1-d}$.

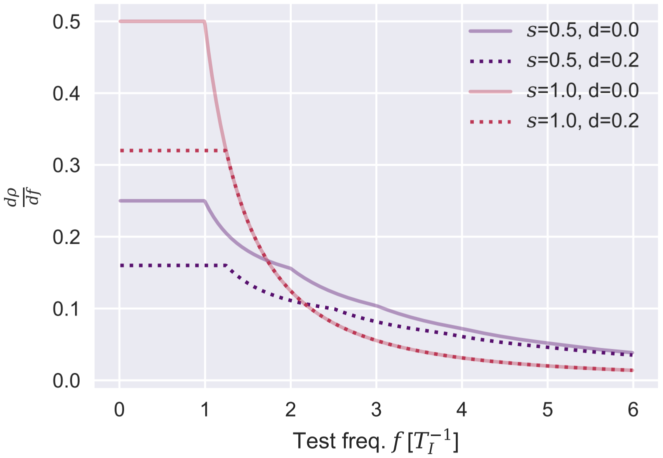


FIG. S2. The derivative $\frac{d}{df}\rho(f, s)$ for sensitivities $s = 1$ and $s = 0.5$ and delays $d = 0$ and $d = 0.2$.
